# Model Evaluation of Secondary Chemistry due to Disinfection of Indoor Air with Germicidal Ultraviolet Lamps

**DOI:** 10.1101/2022.08.25.22279238

**Authors:** Zhe Peng, Shelly L. Miller, Jose L. Jimenez

## Abstract

Air disinfection using Germicidal Ultraviolet light (GUV) has received increasing attention during the COVID-19 pandemic. GUV uses UVC lamps to inactivate microorganisms, but it also initiates photochemistry in air. However, GUV’s indoor-air-quality impact has not been investigated in detail. Here, we model the chemistry initiated by GUV at 254 (“GUV254”) or 222 nm (“GUV222”) in a typical indoor setting for different ventilation levels. Our analysis showed that GUV254, usually installed in the upper room, can significantly photolyze O_3_, generating OH radicals that oxidize indoor volatile organic compounds (VOCs) into more oxidized VOCs. Secondary organic aerosol (SOA) is also formed as a VOC-oxidation product. GUV254-induced SOA formation is of the order of 0.1-1 μg/m^3^ for the cases studied here. GUV222 (described by some as harmless to humans and thus applicable for the whole room) with the same effective virus-removal rate makes a smaller indoor-air-quality impact at mid-to-high ventilation rates. This is mainly because of the lower UV irradiance needed and also less efficient OH-generating O_3_ photolysis than GUV254. GUV222 has a higher impact than GUV254 under poor ventilation due to a small but significant photochemical production of O_3_ at 222 nm, which does not occur with GUV254.

**Synopsis:** Germicidal ultraviolet light initiates indoor oxidation chemistry, potentially forming indoor air pollutants. The amount is not negligible and depends on both the wavelength of light and the ventilation level.

## Introduction

Germicidal ultraviolet light (GUV) has been employed to disinfect air in indoor spaces since the 1930s.^1^ It has been shown to effectively limit the airborne transmission of infectious diseases, e.g., measles and tuberculosis.^1–3^ This is due to photon-induced dimerization of pyrimidines in the nucleic acids of airborne pathogens (and loss of their ability to replicate as a result). GUV fixtures use lamps that emit in the UVC range, most commonly at 254 nm (hereinafter “GUV254”).^4^ As 254 nm UV can cause skin and eye irritation,^5^ GUV254 is usually applied near the ceiling (Figure 1a) or inside ventilation ducts. Recently, 222 nm UV has been shown to not only have strong capability of inactivating airborne viruses,^6^ but also is claimed to be safe to humans^7^ (although this is controversial),^8^ potentially allowing whole-room GUV applications (“GUV222”) (Fig. 1b). Ground-resting GUV-based air cleaners have also been commercialized, in which a fan continuously pulls air into a box and exposes it to UV light, from which the occupants are shielded.^9^

**Figure 1.**
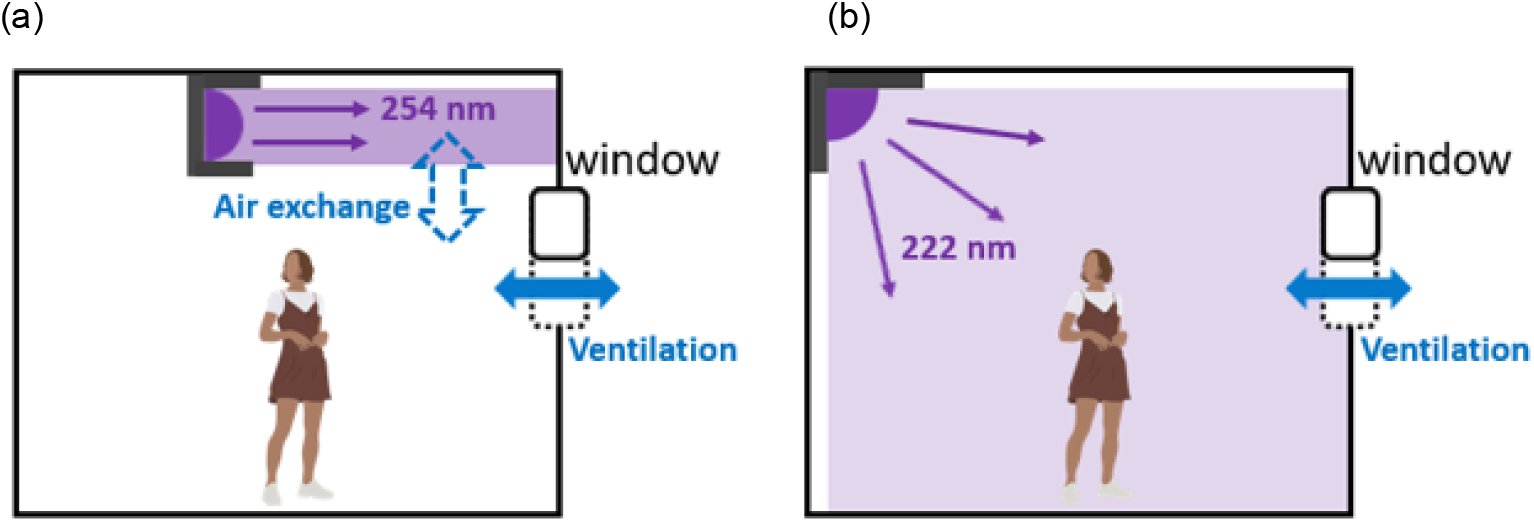
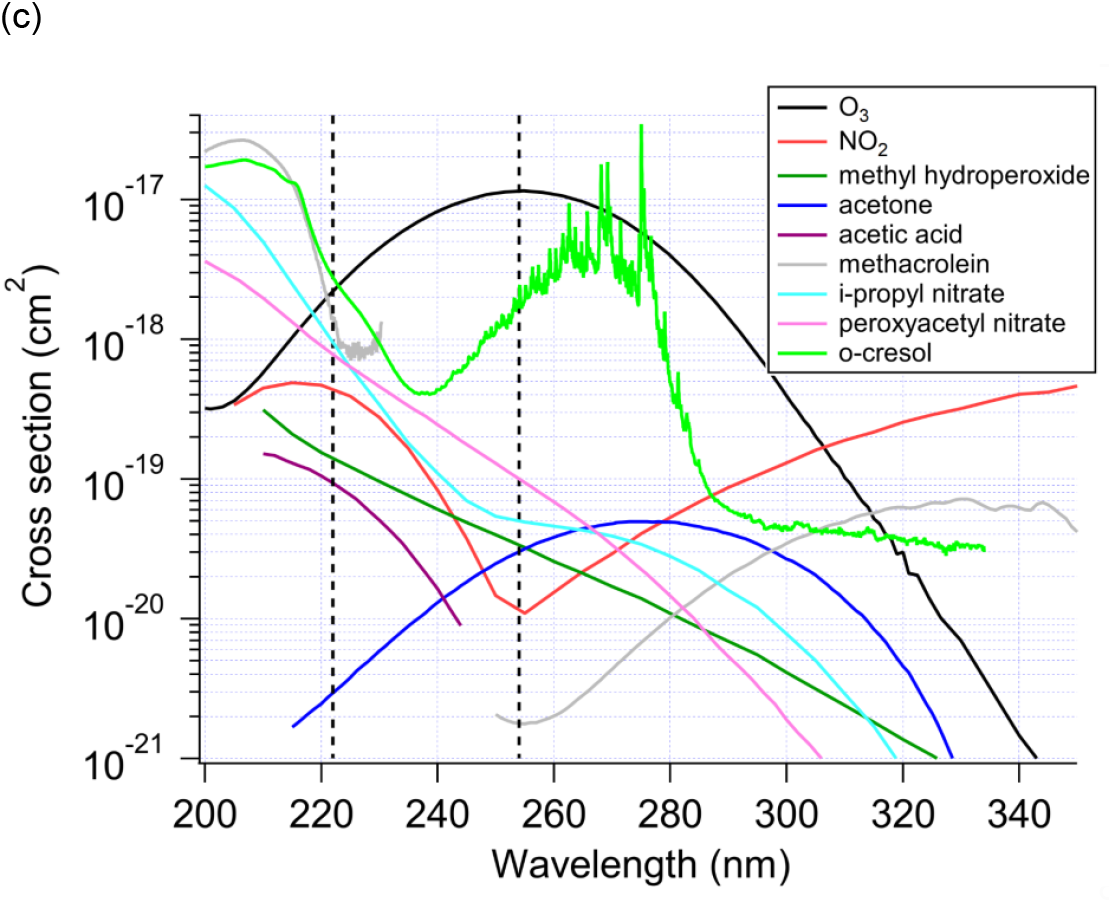
Schematics of a germicidal ultraviolet air disinfection system at (a) 254 nm and at (b) 222 nm in a room; (c) absorption cross sections of several important gas-phase species relevant to this study (a discontinuity in the spectrum of methacrolein is due to lack of data).

During the COVID-19 pandemic, GUV has drawn renewed interest as a tool for airborne virus inactivation. Inhalation of airborne virus is widely accepted as the main transmission route of COVID-19,^10–12^ which explains the dominant indoor character of transmission.^13^ An important component of the transmission is due to superspreading events,^14^ which have been shown to be mostly explained by shared-room airborne transmission.^15^ Much transmission also happens in close proximity due to short-range airborne transmission, but even in this situation a substantial fraction of the inhaled virus may come from well-mixed room air.^16,17^ As COVID-19 remains widespread, and with the possible appearance of new variants, there is a pressing need to remove inhalable viruses from indoor environments.^3^ Similar measures would be beneficial for other airborne diseases such as tuberculosis, measles, or a future pandemic virus.

Physical measures such as (natural and/or mechanical) ventilation and air filtration have been proven safe and effective.^18^ Nevertheless, mechanical ventilation and air filtration usually can remove airborne pathogens only at a few effective air changes per hour (ACH)^19^ and natural ventilation can be highly variable and impractical depending on weather, or when pollution, allergens or noise are present outdoors. When a high virus removal rate (e.g., >10 ACH) needs to be ensured (e.g., in high-risk environments), GUV emerges as a practical and potentially cost-effective way to achieve it.^3,18^

UVC light is known to generate strong oxidants (e.g., OH radicals, and sometimes also O_3_ depending on the wavelengths used),^20^ which can subsequently oxidize volatile organic compounds (VOCs) indoors and initiate organic radical chemistry in indoor air.^21,22^ Energetic UVC photons can also directly photolyze many VOCs, such as peroxides^23,24^ and carbonyls,^25,26^ and generate organic radicals. This radical chemistry is thought to lead to further oxidation of indoor VOCs and the formation of oxygenated VOCs (OVOCs) and secondary organic aerosol (SOA), both of which may have negative health effects.^27^ Surveys of the concentration of total VOCs in the indoor environments range from ∼0.1-4 mg m^-3^.^28–30^ Thus there is always a significant amount of VOC to react with any radicals and oxidants that are generated indoors, and any “air cleaning” technique that can create radicals and/or oxidants indoors has the potential to lead to secondary chemistry.^27^ Very few studies on this topic have been conducted with state-of-the-art measurements or models. Recently, air cleaning devices based on chemistry induced by UV light (photocatalysis and OH generation, but not GUV), often also marketed as suitable for air disinfection, have been experimentally shown to produce significant amounts of OVOCs and SOA^.31,32^

Despite the potential of GUV to cause secondary chemistry, to our knowledge this topic has not been studied in detail to date. Some studies of GUV inactivation effectiveness have included measurements of ozone, to assess whether any was generated.^9^ These studies report no production of ozone when mercury vapor lamps coated to limit emission from wavelengths nearer to the ozone generating wavelength of 185 nm are used, as expected. However, some uncoated or improperly-coated lamps are commercially available, so ozone production can be a problem in some cases. In this study, we perform a first modeling evaluation of the impacts of GUV254 (assuming properly-coated lamps) and GUV222 on indoor air quality. The amounts of OVOC and SOA that can be formed in typical indoor environments are investigated.

## Materials and Methods

We include the photochemistry due to GUV and subsequent radical, oxidation, and SOA formation chemistries. Given the complexity of the composition of indoor air, we simplify both the chemical species present indoors and the reaction scheme, while keeping them consistent with the state-of-the-art knowledge for indoor air. Surface reactions are neglected but could be important, and should be investigated in future studies.

The chemical mechanism for this study is a combination of the inorganic radical chemistry in an oxidation flow reactor (OFR) model^20,33,34^ and part of the Regional Atmospheric Chemistry Mechanism (RACM)^35^ relevant to this study. Section S1 (Supp. Info.) provides more details of the mechanism. The mechanisms are run within the open-source KinSim chemical kinetics simulator,^36^ and are made available (See SI). We perform all simulations until a steady state is reached.

We investigate a typical indoor space with representative indoor and urban outdoor air concentrations from the literature (Table S2). The initial concentrations of most VOCs are estimated based on McDonald et al,^37^ with the total VOC concentration assumed to be 1.7 mg m^-3^, a typical value for US indoor spaces.^28^ See Section S2 for details on species lumping and initial conditions.

The GUV254 fixture in our simulations is based on the AeroMed LEXUS L2.1 Open.^38^ The space irradiated by this device is 45 m^3^, and is placed in a room of 300 m^3^ (volume of a typical classroom), thus the irradiated volume is 15% of the volume of the room, consistent with refs 39,40. The GUV254 model has two compartments, one for the irradiated zone and the other for the rest of the room, while that for GUV222 has only one compartment. Based on the UV inactivation rate constants at 222 nm for SARS-CoV-2,^6,7^ the UV intensity for GUV222 is adjusted such that it provides the same whole-room effective virus-removal rate as GUV254 (see Section S3). Three levels of ventilation, i.e., a representative residential level (0.3 ACH, “low ventilation”),^41^ a representative commercial level (3 ACH, “medium ventilation”),^19^ and a representative medical level (9 ACH, “high ventilation”)^3^ are simulated in this study. Indoor VOC emissions are set such that all VOC concentrations remain at their literature-constrained initial values at low ventilation without chemistry occurring. To test COVID-19 infection risk in different situations, we assume the presence of an infector shedding aerosolized SARS-CoV-2 at 16 quanta h^-1^ (roughly for light exercise while speaking 50% time),^42^ which is consistent or lower than values constrained for literature superspreading events.^15^ A quantum is an infectious dose, that if inhaled by a susceptible person, will lead to a probability of infection of 1-1/e.^15,43^ The rate of SARS-CoV-2 loss apart from ventilation and GUV (i.e., due to intrinsic loss of infectivity, aerosol deposition etc.) is assumed to be 1 h^-1^.

## Results and Discussion

### Disinfection

Figure 2 shows the amount of SARS-CoV-2 present in the room to be consistent with the steady-state prediction. In the absence of GUV, the emission rate of SARS-CoV-2 is 16 quanta h^-1^, and its total loss rate 1.3 h^-1^ (0.3 h^-1^ from ventilation and 1 h^-1^ from decay and deposition) for the low-ventilation case. The steady state SARS-CoV-2 quantity in the entire room is 12.3 quanta. It is lowered to 4 and 1.6 quanta by increasing the ventilation rate to 3 and 9 ACH, respectively. When GUV with a whole-room virus-removal rate of ∼30 h^-1^ is applied, SARS-CoV-2 decreases to ∼0.6, ∼0.5, and ∼0.4 quanta in the low-, medium-, and high-ventilation cases, respectively. The relative impact of GUV is higher at low ventilation, as expected.

**Figure 2.**
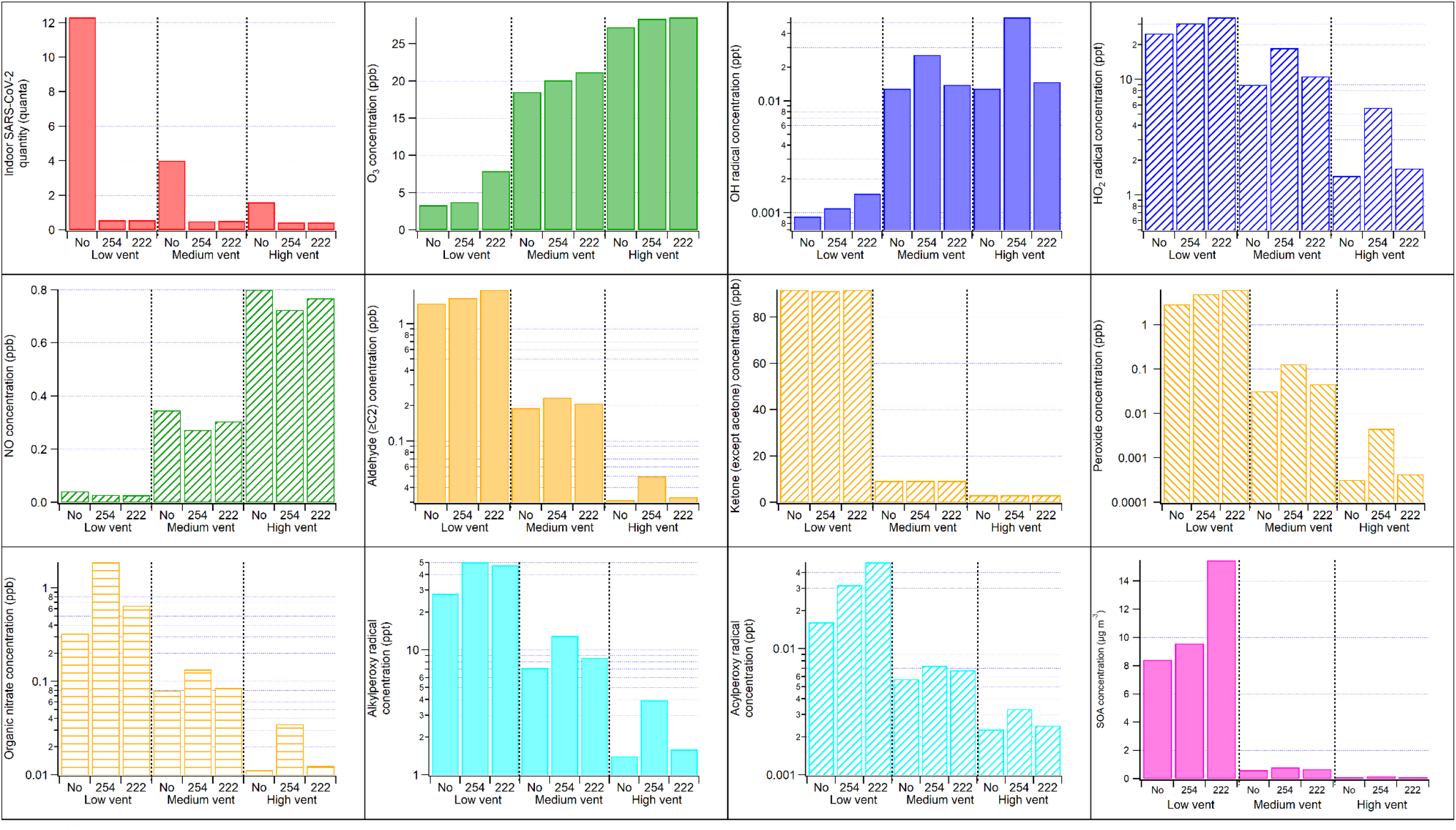
Final quantity/concentration of the main (types of) species of interest in this study under different GUV and ventilation conditions. In the GUV254 cases, the volume-weighted average concentrations for the whole room are shown. The stable chemical species concentrations are similar between the irradiated and unirradiated zones, while the radical and SARS-CoV-2 concentrations in the unirradiated zone can be significantly lower and higher, respectively (Table S3). Note that some panels use log scale for concentrations while other panels use linear scale. SOA is assumed to have a molar weight of 200 g mol^-1^.

The total quantity of SARS-CoV-2 does not directly reflect its infection risk, which also depends on the volume of the room and the inhalation by susceptible occupants. For an occupant with a breathing rate of 0.5 m^3^ h^-1^ (typical for light physical activities)^44^ present in the 300 m^3^ room with low ventilation and no GUV fixture for 1 h, ∼0.02 quantum is inhaled. This corresponds to an infection probability of ∼2%, since the infection probability is approximately equal to the inhaled quanta if the latter is small.^45^ For the cases studied in this work, infection risk is reduced by x∼3 by medium ventilation, and by a factor of ∼22/8/4 when adding GUV to a low/medium/high ventilation situation.

### Secondary Chemistry

For chemical species in the room, ventilation alone (without GUV) can make some difference (Fig. 2). The differences in O_3_, NO, and ketone concentrations are largely due to these species being ventilated in or out. For other chemical species, secondary chemical processes also play a role. OH radicals can form even without UV, i.e., from limonene ozonolysis. As a result, OH radicals are higher at medium and high ventilation, which introduces more O_3_ from outdoors than at low ventilation. OH concentration at high ventilation is not higher than medium ventilation because high ventilation also decreases indoor limonene, reducing the overall limonene-O_3_ reaction rate. HO_2_ radicals are lower at higher ventilation because of higher NO being ventilated into the room, which reacts with HO_2_. All other organic radicals and stable products shown in Fig. 2 (including SOA) have higher concentrations in the low-ventilation case due to higher VOC concentrations.

With GUV254, although the concentration of photolyzable O_3_ remains relatively stable (due to much stronger replenishment from outdoor air ventilation than photolytic destruction), the chemistry is significantly altered by UV (Fig. 2). The fundamental cause of this change is OH production from O_3_ photolysis.^20^ OH concentrations in the GUV254 cases are approximately a factor of 1.2-5 and 3-20 times those in the corresponding no-UV cases for the whole room average and for the irradiated zone, respectively. The difference in the higher-ventilation cases is larger, due to more O_3_ in the room from outside air (Fig. 2 and Table S3). OH in the higher-ventilation cases is similar to daytime outdoor urban levels.^46^ This OH level is high enough to drive substantial oxidation of VOCs, production of other radicals (e.g., HO_2_ and organic peroxy radicals (RO_2_)), and SOA formation. Organic peroxides (including hydroperoxides), carbonyls (aldehydes (excluding formaldehyde) and ketones (excluding acetone)), and organic nitrates (including peroxynitrates) are among common VOC oxidation products and all have ∼10% to several-fold concentration increases (Fig. 2). The exceptions are ketones (excluding acetone), whose production is dominated in the model by UV-independent limonene ozonolysis (Fig. 2). Doubling RH significantly increases OH concentration, but changes of product species are within 10% of the base RH results, likely due to non-linear buffering from the reaction scheme (not shown). The exceptions are ketones, which are relatively unreactive and dominated in the model by the chemistry-independent acetone emission and its dilution by ventilation (Fig. 2). In addition to VOC oxidation by OH, radicals (OH, HO_2_, and RO_2_) are also produced by active photolysis of carbonyls and peroxides at 254 nm, where both strongly absorb (Fig. 1c). Due to higher peroxy radical concentrations, NO is lowered to ∼30 ppt at low ventilation (Fig. 2). Such a low NO concentration leads to reactions of RO_2_ with both HO_2_ and NO being important, as estimated per ref 47 (Section S4).

SOA formation is estimated from the consumption of individual VOCs and SOA mass yields from the literature (Table S4). Significant SOA production (∼8 μg m^-3^ at low ventilation) occurs even without GUV irradiation, through limonene ozonolysis, as this reaction has a high SOA yield (20%). In the GUV254 cases, both limonene ozonolysis and VOC oxidation by OH contribute to SOA. The overall SOA mass yield from total VOC is of the order of 0.1%, because a large fraction of total organic molecules present are too small to form SOA through oxidation (e.g., ethanol or acetone). Given that 1.5 mg m^-3^ of total VOCs (excluding limonene) are present, that leads to ∼1.2 μg m^-3^ due to GUV254. The enhancement of SOA formation by OH oxidation relative to the no-UV cases decreases with increasing ventilation, as VOC concentrations are lowered by ventilation (Fig. 2). The SOA precursors and mechanism used in this work are likely incomplete, given the incomplete scientific understanding of this topic.^48^

Due to the fast air exchange between the GUV254 irradiated and unirradiated spaces, the concentrations of stable species are similar between these two spaces (Table S3). In contrast, radicals are more rapidly consumed in the unirradiated space than supplied by the transport from the irradiated space, and thus have much lower concentrations (up to >1 order of magnitude for highly reactive ones such as OH and acylperoxy) in the unirradiated space.

The GUV222 cases assume irradiation of the entire room volume (Fig. 1b). 222 nm photons can photolyze O_2_ and produce O_3_, albeit at a small rate, leading to higher O_3_ in all cases relative to GUV254. The amounts of organic products formed in the medium- and high-ventilation (low-ventilation) cases are lower (higher) than in the unirradiated zone in the GUV254 cases (Fig. 2).

At medium and high ventilation rates, the main O_3_ source in the GUV222 cases is still outdoor O_3_ through ventilation. Despite some O_3_ production, the product enhancement is much smaller than in the corresponding GUV254 cases (Fig. 2). These results indicate a weak OH-initiated VOC oxidation on top of the VOC ozonolysis chemistry that is active in the no-UV cases.

This difference from the active photochemistry in the GUV254 cases can be attributed to several factors. First, the UV irradiance of the 222 nm fixture is significantly lower, even in terms of the number of photons emitted per unit time. 222 nm photons are ∼40% more efficient in inactivating SARS-CoV-2 than 254 nm photons.^6^ In addition the latter cannot be used in the most efficient fashion. Due to the need to protect humans from irradiation, all photons are concentrated in the small irradiated zone (15% of the room volume). Because of the limited rate of transport of virus-containing aerosol to the irradiated zone, the steady-state infectious virus concentration is ∼70% lower than in the unirradiated zone, where the infector and the susceptible individuals are present (Table S3). Even if the per-photon virus-inactivation efficiency was the same, GUV254 would need about 3 times the photons for GUV222 to reach the same effective GUV virus-removal rate for the occupied unirradiated space. Furthermore, the first step of OH photochemical production is O_3_ photolysis, whose corresponding absorption at 222 nm is about ∼5 times lower than at 254 nm (Fig. 1c). Simple carbonyl compounds, the most abundant OVOCs in this study, also absorb much less efficiently at 222 nm than at 254 nm (Fig. 1c), further reducing radical production. Although other products, such as peroxides and conjugated carbonyl species, can have stronger absorption at 222 nm, their relatively low concentrations (∼1 ppb or lower vs. hundreds of ppb of ketones) limit their relative contributions to the radical budget.

Because of the small direct production of O_3_ by the GUV222 lights, O_3_ in the GUV222 low-ventilation case is substantially less depleted by limonene ozonolysis than for GUV254. As a result, compared to GUV254 SOA formation through limonene ozonolysis is substantially stronger and OH concentration is also higher (Fig. 2). Other gas-phase stable organic products have comparable concentrations to those in the GUV254 case.

### Implications

We have shown that GUV disinfection can induce active photochemistry producing OVOCs and SOA in typical indoor environments. Under the conditions simulated here, these products do not necessarily have significant negative effects on human health because of their relatively low concentrations. Among the VOCs (including OVOCs) modeled in this study, only formaldehyde has a concentration exceeding the Minimal Risk Level (MRL) recommended by the CDC^49^ due to strong indoor emissions. However, only a very limited number of species were explicitly modeled in this study, particularly aldehydes, whose toxicity is generally high. Future studies with higher chemical speciation are needed to better assess the toxicity of gas-phase products. In polluted indoor spaces and/or outdoor atmospheric environments, the indoor concentrations of the VOCs of interest can be much higher,^28^ while the GUV-induced photochemistry can still be active (Sections S5-S6). In this case, OVOC products might exceed the MRLs and SOA formation might reach tens of μg m^-3^.

The risk of GUV254 due to secondary photochemical products is not negligible but also not dominant under typical indoor conditions. The risk for GUV222 appears to be substantially lower when ventilation is not poor, but comparable or slightly higher in case of poor ventilation. We note that many indoor environments, in particular homes and schools, have ventilation rates similar to the definition of “poor” in this paper, even in high-income countries. If GUV222 is confirmed to be safe for direct human exposure, it would have an advantage over GUV254 at mid to high ventilation rates in terms of indoor chemistry, in addition to more efficient air disinfection. When GUV254 is used, a strong air exchange between the irradiated and unirradiated zones (e.g., by fans) is preferable, as recommended by the CDC/NIOSH.^50^ It can lower the UV irradiance needed for a given virus inactivation rate,^51^ and hence limit the induced photochemistry. Good ventilation can not only remove airborne pathogens, but also limit the production of secondary indoor pollutants, and is thus also recommended when outdoor air is relatively clean.^19,52^ Similarly, particulate air filtration is also recommended as it removes both virus-containing aerosol, indoor-formed SOA, and particulate pollution from other indoor and outdoor sources. Gas filtration with sorbent materials such as activated carbon is also useful for reducing VOC, NO_x_ and ozone levels indoors.^53,54^ The findings of this study are limited due to modeling assumptions, e.g., the simplified chemical mechanism, limited data on photolysis parameters in the UVC range, the assumed indoor and outdoor air pollutant compositions, uncertainties over precursors and yields of SOA formation, and surface reactions. Experimental studies in both simplified laboratory settings and real indoor conditions are needed to fully constrain the impacts of GUV in indoor chemistry.

## Data Availability

All data produced in the present study are available upon reasonable request to the authors.

## Acknowledgements

ZP and JLJ were supported by the CIRES Innovative Research Program and the Balvi Filantropic Fund. We thank Vito Ilacqua, Zachary Finewax, Donald Milton, and Edward Nardell for valuable discussions.

## Associated Content

This paper has been previously submitted to medRxiv, a preprint server for health sciences. The preprint can be cited as Peng, Z.; Miller, S. L.; Jimenez, J. L. Model Evaluation of Secondary Chemistry due to Disinfection of Indoor Air with Germicidal Ultraviolet Lamps. 2022. medRxiv 10.1101/2022.08.25.22279238v3 (accessed November 16, 2022).

## Supporting Information

The Supporting Information is available free of charge at https://pubs.acs.org/doi/10.1021/acs.estlett.xxxxxxx.

Details about the model setup (reaction scheme, species lumping, indoor emission, outdoor air composition, initial conditions, effective virus-removal rates of GUV), RO_2_ fates in the GUV254 cases, concentrations in the irradiated and unirradiated zones in the GUV254 cases, and sensitivity cases for highly polluted conditions.

