## Supporting Information for "Model Evaluation of Secondary Chemistry due to Disinfection of Indoor Air with Germicidal Ultraviolet Lamps"

**S1. Oxidation flow reactor mechanism and part of the Regional Atmospheric Chemistry Mechanism (RACM) used in this study**

The OFR employs UVC lamps, with the explicit purpose of generating radicals that initiate oxidation reactions. OFR are used extensively in atmospheric chemistry research. A common OFR operation mode ("OFR254") uses 254 nm UV light from filtered mercury lamps to photolyze $O_3$, through which $O(^1D)$ is generated, which subsequently reacts with water vapor to form OH.[1,2] OFR254 uses the same type of lamps as GUV254 and thus OH is expected to form through the same chemistry. Therefore, all reactions of this inorganic radical chemistry are adopted.[2]

For organic chemistry, the relevant reactions in RACM are adopted. Among major relevant (lumped) species are HC8 (alkanes, alcohols, and esters with relatively fast reaction rate with OH), LIM (limonene), KET (ketones), ALD (aldehydes), OP2 (higher organic peroxides), and ACO3 (acylperoxy radicals). As all inorganic reactions are explicitly included in the OFR mechanism, which is more suitable for the chemistry under GUV, the inorganic part of the RACM mechanism is not used in this study. For simplicity, we generally only include the organic species that are emitted in the case studies in the present work and those that are products of the emitted species in the RACM. Products of a reaction with a yield (stoichiometric coefficient) <0.1 are considered minor and neglected in that reaction. A few reactions (e.g., photolysis of acetic acid (Fig. 1b)) are added to include the chemistry that can occur under UVC irradiation but cannot under UVA and UVB irradiation (only the latter is accounted for by the RACM). All photolysis cross sections are adopted from refs 3,4 when available, otherwise estimated from those of molecules containing the same functional groups according to the framework of Peng et al.[5] All quantum efficiencies for reactant photodissociation except those with available data in refs 3,4 are assumed to be 1, given the high photon energies involved. For GUV222, the photolysis frequencies are calculated using the light flux at that wavelength. Also, as $NO_3$ concentration is very low in this study, reactions of organics with $NO_3$ are neglected. All organic reactions used in this study are listed in Table S1.

**S2. Lumping of the species in the emission inventory of McDonald et al. into species used in the RACM and initial conditions of the model cases**

We lump the (indoor fraction of the) species listed in Table S8 of McDonald et al.[6] into the most similar species in the RACM. The mass fractions of the species in McDonald et al.'s emission inventory in the total VOC are assigned to the mass fractions of the corresponding RACM species. The indoor emission fractions of the RACM species are estimated as a rough average of those of the lumped species in the McDonald et al. inventory. The resulting indoor mass fractions of the RACM species are renormalized, and with a total VOC of 1.7 mg $m^{-3}$ assumed, the concentrations of individual RACM species can be calculated. The correspondence of the RACM species to those in the McDonald et al. inventory and the estimated indoor emission fractions and normalized mass fractions of the RACM species are shown in Table S2. The only exceptions to this initial VOC concentration estimation are that we assign higher concentrations of 300 ppb to acetone as measured by Price et al.[7] and of 2 ppm to formaldehyde per ref 8.

The atmospheric pressure and temperature in the room are assumed to be 1 atm and 295 K, respectively, with a relative humidity of 37% (water vapor mixing ratio of 1%), and initial NO, HONO, $NO_2$, and $O_3$ concentrations of 1, 5, 10, and 10 ppbv, respectively. We assume no $NO_x$ or $O_3$ emissions indoors, and 5 ppb NO, 20 ppb $NO_2$, 40 ppb $O_3$ outdoors. No VOC is assumed to be present in outdoor air, as outdoor VOC levels are typically much lower than indoor ones.[7,9] The $O_3$ surface loss rate in the absence of chemistry is set to 2.8 $h^{-1}$, which is typical of residences[10] and is known to be sensitive to occupancy and the indoor surfaces present.

**S3. Indoor air exchange rate for the GUV254 case and UV intensity for the GUV222 fixture**

The GUV254 fixture in this study has a UV intensity of ~$1 \times 10^{14}$ photons $cm^{-2}$ $s^{-1}$ for the averaged irradiated section area of 4.5 $m^2$, leading to a virus inactivation rate of ~500 $h^{-1}$ for the irradiated zone. We set an air exchange rate for the irradiated zone at 240 $h^{-1}$ (with the unirradiated zone).[11] As the volume of the irradiated space is 15% of the whole room, the effective virus removal rate for the whole room by GUV254 is ~30 $h^{-1}$ (equivalent ACH) using a SARS-CoV-2 UV inactivation rate of 0.79 $cm^2$/mJ.[12] Such an equivalent ACH is representative of well-designed indoor GUV applications.[13–15] Air within a modeled indoor air compartment is assumed to be well mixed. Although highly reactive radicals (e.g., OH) may not travel far from the irradiated zone due to short lifetimes,[16] their concentrations can still be averaged over the entire unirradiated space because of their low concentrations and thus the low importance of self- and cross-reactions in their fates.

For the 222 nm UV, we assume its intensity to be uniform all over the room. The same effective virus removal rate as in the occupied unirradiated zone for GUV254 corresponds to a UV intensity of $3.9 \times 10^{12}$ photons $cm^{-2}$ $s^{-1}$, with a UV inactivation rate coefficient of 1.42 $cm^2$ $mJ^{-1}$ for SARS-CoV-2 at 222 nm.[12]

**S4. Organic peroxy radical fates in the GUV254 cases**

Due to low NO concentration (~30 ppt) at low ventilation, $RO_2$+NO and $RO_2$+$HO_2$ both account for nearly half of the $RO_2$ bimolecular loss in the irradiated space (Fig. S1). Also, without fast $RO_2$+NO, $RO_2$ lifetime is sufficiently long for unimolecular reactions of $RO_2$ to occur (Fig. S1) as observed previously indoors for similar conditions,[17] although RACM does not include these reactions. In the higher ventilation cases, NO, though still consumed by the photochemistry, is much higher due to a stronger replenishment from outdoor air and can dominate the $RO_2$ bimolecular fate (Fig. S1).

**S5. Sensitivity GUV cases with high VOC emissions**

These sensitivity cases for GUV254 and GUV222 have x10 indoor VOC emissions as an example of a very polluted indoor space. All other settings for this case are the same as the corresponding GUV254 and GUV222 cases with low ventilation. Compared to the standard low-ventilation GUV cases, the steady-state concentrations of gas-phase organic products and SOA are ~5-10 times higher. $O_3$ is lower because of stronger limonene ozonolysis. Relative to a no-UV case with x10 VOC emissions, SOA formation enhancement due to GUV is ~4 μg $m^{-3}$ in the GUV254 sensitivity case. In the GUV222 sensitivity case, GUV-enhanced SOA formation is ~60 μg $m^{-3}$, largely due to limonene ozonolysis promoted by GUV222-induced $O_3$ production. These SOA enhancements account for similar fractions of total SOA formation as in the standard low-ventilation GUV254 and GUV222 cases.

**S6. Sensitivity GUV cases with wildfire smoke in the outdoor air**

These sensitivity cases for GUV254 and GUV222 have a highly polluted outdoor air composition. We assumed that in the outdoor air there is wildfire smoke with an OA concentration of 200 μg $m^{-3}$, which is an upper limit for wildfire smoke in urban areas in the Western US.[18] The fractions of gas-phase organic species in the total organic carbon are estimated based on the wildfire plume organic species concentrations reported by Heald et al.[19] These species are lumped with the most similar species in the chemical mechanism used in this study. The NO concentration is estimated based on the emission factors reported by Urbanski.[20]

Compared to the base low-ventilation GUV254 and GUV222 cases, all changes in the stable organic product concentrations are within 20%, except for those with high concentrations outdoors, e.g., OA and PAN. The increases in the concentrations of these species are dominantly due to ventilation. At higher ventilation rates, the increases in the indoor concentrations of even more species (e.g., NO and ketones) are dominated by the ventilation.

(13) Xu, P.; Kujundzic, E.; Peccia, J.; Schafer, M. P.; Moss, G.; Hernandez, M.; Miller, S. L. Impact of Environmental Factors on Efficacy of Upper-Room Air Ultraviolet Germicidal

Irradiation for Inactivating Airborne Mycobacteria. *Environ. Sci. Technol.* **2005**, *39* (24), 9656–9664.
(14) Xu, P.; Peccia, J.; Fabian, P.; Martyny, J. W.; Fennelly, K. P.; Hernandez, M.; Miller, S. L. Efficacy of Ultraviolet Germicidal Irradiation of Upper-Room Air in Inactivating Airborne Bacterial Spores and Mycobacteria in Full-Scale Studies. *Atmos. Environ.* **2003**, *37* (3), 405–419.
(15) Nardell, E. A. Air Disinfection for Airborne Infection Control with a Focus on COVID‐19: Why Germicidal UV Is Essential. *Photochem. Photobiol.* **2021**, *97* (3), 493–497.
(16) Lakey, P. S. J.; Won, Y.; Shaw, D.; Østerstrøm, F. F.; Mattila, J.; Reidy, E.; Bottorff, B.; Rosales, C.; Wang, C.; Ampollini, L.; Zhou, S.; Novoselac, A.; Kahan, T. F.; DeCarlo, P. F.; Abbatt, J. P. D.; Stevens, P. S.; Farmer, D. K.; Carslaw, N.; Rim, D.; Shiraiwa, M. Spatial and Temporal Scales of Variability for Indoor Air Constituents. *Communications Chemistry* **2021**, *4* (1), 110.
(17) Pagonis, D.; Algrim, L. B.; Price, D. J.; Day, D. A.; Handschy, A. V.; Stark, H.; Miller, S. L.; de Gouw, J. A.; Jimenez, J. L.; Ziemann, P. J. Autoxidation of Limonene Emitted in a University Art Museum. *Environ. Sci. Technol. Lett.* **2019**, *6* (9), 520–524.
(18) O'Dell, K.; Ford, B.; Burkhardt, J.; Magzamen, S.; Anenberg, S. C.; Bayham, J.; Fischer, E. V.; Pierce, J. R. Outside in: The Relationship between Indoor and Outdoor Particulate Air Quality during Wildfire Smoke Events in Western US Cities. *Environ. Res.: Health* **2022**. https://doi.org/10.1088/2752-5309/ac7d69.
(19) Heald, C. L.; Goldstein, A. H.; Allan, J. D.; Aiken, A. C.; Apel, E.; Atlas, E. L.; Baker, A. K.; Bates, T. S.; Beyersdorf, A. J.; Blake, D. R.; Others. Total Observed Organic Carbon (TOOC) in the Atmosphere: A Synthesis of North American Observations. *Atmos. Chem. Phys.* **2008**, *8* (7), 2007–2025.
(20) Urbanski, S. Wildland Fire Emissions, Carbon, and Climate: Emission Factors. *For. Ecol. Manage.* **2014**, *317*, 51–60.
(21) Stockwell, W. R.; Kirchner, F.; Kuhn, M.; Seefeld, S. A New Mechanism for Regional Atmospheric Chemistry Modeling. *J. Geophys. Res.* **1997**, *102* (D22), 25847–25879.
(22) Peng, Z.; Jimenez, J. L. KinSim: A Research-Grade, User-Friendly, Visual Kinetics Simulator for Chemical-Kinetics and Environmental-Chemistry Teaching. *J. Chem. Educ.* **2019**, *96* (4), 806–811.
(23) Saathoff, H.; Naumann, K.-H.; Möhler, O.; Jonsson, Å. M.; Hallquist, M.; Kiendler-Scharr, A.; Mentel, T. F.; Tillmann, R.; Schurath, U. Temperature Dependence of Yields of Secondary Organic Aerosols from the Ozonolysis of α-Pinene and Limonene. *Atmos. Chem. Phys.* **2009**, *9* (5), 1551–1577.
(24) Peng, Z.; Lee-Taylor, J.; Orlando, J. J.; Tyndall, G. S.; Jimenez, J. L. Organic Peroxy Radical Chemistry in Oxidation Flow Reactors and Environmental Chambers and Their Atmospheric Relevance. *Atmos. Chem. Phys.* **2019**, *19* (2), 813–834.
(25) Praske, E.; Otkjær, R. V.; Crounse, J. D.; Hethcox, J. C.; Stoltz, B. M.; Kjaergaard, H. G.; Wennberg, P. O. Atmospheric Autoxidation Is Increasingly Important in Urban and Suburban North America. *Proceedings of the National Academy of Sciences* **2018**, *115* (1), 64–69.
(26) Orlando, J. J.; Tyndall, G. S. Laboratory Studies of Organic Peroxy Radical Chemistry: An Overview with Emphasis on Recent Issues of Atmospheric Significance. *Chem. Soc. Rev.* **2012**, *41* (19), 6294–6317.

**Table S1.** Organic (a) thermal and (b) photolytic reactions in the chemical scheme in this study. See Table 1 of Stockwell et al.[21] for the species names that are not common names or chemical formulas. Note that some reactions with the same reactants are divided into multiple reactions to fit the 3-product requirement of KinSim.[22]

(a)

| Reactants | | Products | | | Rate coefficient ($cm^3$ molecules$^{-1}$ $s^{-1}$ or $s^{-1}$) |
|---|---|---|---|---|---|
| CH3OO | HO2 | CH3OOH | | | 5.20E-12 |
| CH3OO | NO | HCHO | NO2 | | 7.60E-12 |
| ACO3 | NO | CH3OO | NO2 | | 2.00E-11 |
| CH3OOH | OH | CH3OO | H2O | | 7.40E-12 |
| HCHO | OH | CO | H2O | HO2 | 8.50E-12 |
| CO | OH | CO2 | HO2 | | 2.10E-13 |
| CH3OO | CH3OO | 2 HCHO | 2 HO2 | | 1.23E-13 |
| CH3OO | CH3OO | HCHO | CH3OH | | 2.28E-13 |
| CH3OH | OH | HCHO | HO2 | H2O | 9.10E-13 |
| ACO3 | NO2 | PAN | | | 8.66E-12 |
| ACO3 | HO2 | CH3CO3H | | | 5.60E-12 |
| ACO3 | HO2 | CH3COOH | O3 | | 2.80E-12 |
| ACO3 | HO2 | CH3OO | OH | | 5.60E-12 |
| LIM | OH | LIMP | | | 1.71E-10 |
| LIMP | NO | 1.3 HO2 | 0.8 MACR | 0.5 OLI | 2.00E-12 |
| LIMP | NO | 0.5 HCHO | 0.7 ONIT | 1.3 NO2 | 2.00E-12 |
| LIMP | HO2 | OP2 | | | 1.50E-11 |
| LIMP | CH3OO | 2.8 HCHO | 1.2 MACR | 0.8 OLI | 1.92E-13 |
| LIMP | CH3OO | 4 HO2 | | | 1.92E-13 |

| | | | | | |
|---|---|---|---|---|---|
| LIMP | ACO3 | 1.2 MACR | 0.8 HCHO | 0.8 OLI | 4.82E-12 |
| LIMP | ACO3 | 2 HO2 | 2 CH3OO | | 4.82E-12 |
| MACR | OH | 0.51 ACO3 | 0.41 HKET | 0.49 HO2 | 3.35E-11 |
| MACR | O3 | 1.2 HCHO | 1.8 MGLY | | 3.80E-19 |
| MACR | O3 | 0.39 CH3COOH | 0.66 HCOOH | 0.21 OH | 3.80E-19 |
| MACR | O3 | 0.87 HO2 | 0.39 OP2 | 0.39 ACO3 | 3.80E-19 |
| LIM | O3 | 0.92 OLT | 0.32 ETHP | 0.84 KETP | 1.00E-16 |
| LIM | O3 | 1.7 OH | 0.2 HO2 | 1.58 MACR | 1.00E-16 |
| OLI | OH | OLIP | | | 7.12E-11 |
| OLI | O3 | 1.98 ALD | 0.32 KET | 0.28 CH3COOH | 1.29E-16 |
| OLI | O3 | 0.44 HO2 | 1.26 OH | 0.46 CH3OO | 1.29E-16 |
| OLIP | NO | 2 HO2 | 2 NO2 | | 2.00E-12 |
| OLIP | NO | 3.42 ALD | 0.58 KET | | 2.00E-12 |
| OLIP | HO2 | OP2 | | | 1.00E-11 |
| OLIP | CH3OO | 1.51 HCHO | 2 HO2 | | 4.93E-13 |
| OLIP | CH3OO | 1.864 ALD | 0.626 KET | | 4.93E-13 |
| OLIP | ACO3 | 1.882 ALD | 1.138 KET | 1.02 HO2 | 3.32E-12 |
| OLIP | ACO3 | 1.02 CH3OO | 0.98 CH3COOH | | 3.32E-12 |
| ONIT | OH | HC3P | NO2 | | 2.22E-12 |
| OP2 | OH | 0.44 HC3P | 0.41 KET | 0.49 OH | 6.43E-12 |
| HKET | OH | HO2 | MGLY | | 3.00E-12 |
| MGLY | OH | ACO3 | | | 1.72E-11 |
| OLT | OH | OLTP | | | 3.06E-11 |

| | | | | | |
|---|---|---|---|---|---|
| OLTP | NO | 1.88 ALD | 0.12 KET | | 2.00E-12 |
| OLTP | NO | 2 HCHO | 2 HO2 | 2 NO2 | 2.00E-12 |
| OLTP | HO2 | OP2 | | | 1.30E-11 |
| OLTP | CH3OO | 1.25 HCHO | HO2 | 0.669 ALD | 1.57E-12 |
| ETHP | NO | ALD | HO2 | NO2 | 8.70E-12 |
| ETHP | HO2 | OP2 | | | 7.86E-12 |
| ETHP | CH3OO | 0.75 HCHO | HO2 | 0.75 ALD | 2.01E-13 |
| ETHP | ACO3 | 2 ALD | HO2 | | 1.05E-12 |
| ETHP | ACO3 | CH3OO | CH3COOH | | 1.05E-12 |
| KETP | NO | 1.08 MGLY | 0.92 ALD | 0.46 ACO3 | 2.00E-12 |
| KETP | NO | 1.54 HO2 | 2 NO2 | | 2.00E-12 |
| KETP | HO2 | OP2 | | | 9.02E-12 |
| KETP | CH3OO | 1.5 HCHO | 1.76 HO2 | 0.8 MGLY | 1.90E-12 |
| KETP | CH3OO | 0.6 ALD | 0.6 HKET | 0.24 ACO3 | 1.90E-12 |
| KETP | ACO3 | 1.62 MGLY | 1.05 ALD | 0.33 KET | 1.67E-12 |
| KETP | ACO3 | 0.36 ACO3 | 1.14 HO2 | 1.5 CH3OO | 1.67E-12 |
| KETP | ACO3 | 1.5 CH3COOH | | | 1.67E-12 |
| ALD | OH | ACO3 | | | 1.69E-11 |
| KET | OH | KETP | | | 6.87E-13 |
| HC3P | NO | 0.466 ALD | 1.246 KET | 1.484 HO2 | 2.00E-12 |
| HC3P | NO | 0.3 CH3OO | 1.882 NO2 | | 2.00E-12 |
| HC3P | HO2 | OP2 | | | 1.30E-11 |
| HC3P | CH3OO | 0.81 HCHO | 0.992 HO2 | 0.58 ALD | 4.02E-13 |
| HC3P | ACO3 | 1.448 ALD | 0.254 KET | 0.976 HO2 | 1.62E-12 |

| | | | | | |
|---|---|---|---|---|---|
| HC3P | ACO3 | 1.016 CH3OO | 0.998 CH3COOH | | 1.62E-12 |
| ACO3 | ACO3 | 2 CH3OO | | | 1.66E-11 |
| ACO3 | CH3CO3 | 2 CH3OO | | | 1.66E-11 |
| ACO3 | CH3OO | HCHO | HO2 | CH3OO | 7.33E-12 |
| ACO3 | CH3OO | HCHO | CH3COOH | | 1.22E-12 |
| HC3 | OH | 0.583 HC3P | 0.381 HO2 | 0.335 ALD | 2.20E-12 |
| HC8 | OH | 0.951 HC8P | | | 1.08E-11 |
| HC8P | NO | 0.3 ALD | 1.282 KET | 0.266 ETHP | 2.00E-12 |
| HC8P | NO | 0.522 ONIT | 1.478 NO2 | 1.212 HO2 | 2.00E-12 |
| TOL | OH | TOLP | | | 5.96E-12 |
| XYL | OH | XYLP | | | 2.40E-11 |
| HC8P | HO2 | OP2 | | | 1.30E-11 |
| HC8P | CH3OO | 1.506 HCHO | 1.986 HO2 | | 1.82E-13 |
| HC8P | CH3OO | 0.822 ALD | 0.838 KET | | 1.82E-13 |
| HC8P | ACO3 | 0.994 ALD | 1.162 KET | | 1.22E-12 |
| HC8P | ACO3 | 1.014 CH3OO | 0.99 CH3COOH | | 1.22E-12 |
| TOLP | NO | 1.9 NO2 | 1.9 HO2 | | 2.00E-12 |
| TOLP | NO | 1.3 MGLY | 2.4 GLY | DCB | 2.00E-12 |
| XYLP | NO | 1.9 NO2 | 1.9 HO2 | | 2.00E-12 |
| XYLP | NO | 1.2 MGLY | 0.7 GLY | 1.9 DCB | 2.00E-12 |
| TOLP | HO2 | OP2 | | | 1.01E-11 |
| XYLP | HO2 | OP2 | | | 1.01E-11 |
| TOLP | CH3OO | 2 HCHO | 2 HO2 | 2 DCB | 1.92E-13 |

| | | | | | |
|---|---|---|---|---|---|
| TOLP | CH3OO | 0.7 MGLY | 1.3 GLY | | 1.92E-13 |
| XYLP | CH3OO | 2 HCHO | 2 HO2 | 2 DCB | 1.92E-13 |
| XYLP | CH3OO | 1.26 MGLY | 0.74 GLY | | 1.92E-13 |
| TOLP | ACO3 | 2 CH3OO | 2 HO2 | 2 DCB | 4.82E-12 |
| TOLP | ACO3 | 0.7 MGLY | 1.3 GLY | | 4.82E-12 |
| XYLP | CH3OO | 2 HCHO | 2 HO2 | 2 DCB | 4.82E-12 |
| XYLP | CH3OO | 1.26 MGLY | 0.74 GLY | | 4.82E-12 |
| GLY | OH | HO2 | | | 1.14E-11 |
| CH3CH2OH | OH | ALD | HO2 | | 3.20E-12 |
| iPrOH | OH | acetone | HO2 | | 5.10E-12 |
| PAN | | ACO3 | NO2 | | 0.00033 |

(b)

| Reactants / UV photons | | Products | | | Cross section ($cm^2$) |
|---|---|---|---|---|---|
| O2 | UV at 222 nm | 2 O(3P) | | | 4.09E-24 |
| acetone | | CO | 2 CH3OO | | 1.03E-21 |
| acetone | | ACO3 | CH3OO | | 1.91E-21 |
| CH3OOH | | HCHO | OH | | 1.41E-19 |
| HCHO | | 0.61 HO2 | 0.7 CO | 0.495 H2 | 1.80E-22 |
| PAN | | ACO3 | NO2 | | 6.22E-19 |
| PAN | | CH3OO | NO3 | | 1.55E-19 |
| CH3CO3H | | CH3OO | OH | | 1.60E-19 |
| MACR | | ACO3 | HCHO | HO2 | 1.40E-18 |
| ONIT | | 0.4 ALD | 1.6 KET | | 5.00E-19 |

| | | | | | |
|---|---|---|---|---|---|
| ONIT | | 2 HO2 | 2 NO2 | | 5.00E-19 |
| OP2 | | ALD | HO2 | OH | 1.41E-19 |
| HKET | | HCHO | HO2 | ACO3 | 2.94E-21 |
| MGLY | | HO2 | ACO3 | | 1.43E-20 |
| CH3COOH | | ACO3 | OH | | 5.09E-20 |
| CH3COOH | | CH3OO | HO2 | | 4.34E-20 |
| HCOOH | | OH | HO2 | | 1.24E-19 |
| ALD | | CH3OO | HO2 | | 6.50E-22 |
| KET | | ETHP | ACO3 | | 2.94E-21 |
| GLY | | 0.45 HCHO | 0.8 HO2 | | 6.16E-21 |
| acetone | UV at 254 nm | CO | 2 CH3OO | | 1.06E-20 |
| acetone | | ACO3 | CH3OO | | 1.95E-20 |
| CH3OOH | | HCHO | OH | | 3.40E-20 |
| HCHO | | 0.61 HO2 | 0.7 CO | 0.495 H2 | 3.69E-21 |
| PAN | | ACO3 | NO2 | | 8.00E-20 |
| PAN | | CH3OO | NO3 | | 2.00E-20 |
| CH3CO3H | | CH3OO | OH | | 2.42E-20 |
| MACR | | ACO3 | HCHO | HO2 | 1.79E-21 |
| ONIT | | 0.4 ALD | 1.6 KET | | 2.50E-20 |
| ONIT | | 2 HO2 | 2 NO2 | | 2.50E-20 |
| OP2 | | ALD | HO2 | OH | 3.40E-20 |
| HKET | | HCHO | HO2 | ACO3 | 4.98E-20 |
| MGLY | | HO2 | ACO3 | | 2.76E-20 |

| | | | | | |
|---|---|---|---|---|---|
| ALD | | CH3OO | HO2 | | 1.57E-20 |
| KET | | ETHP | ACO3 | | 3.01E-20 |
| GLY | | 0.45 HCHO | 0.8 HO2 | | 1.41E-20 |

**Table S2.** Correspondences of the RACM species to those in the McDonald et al.[6] emission inventory and estimated indoor emission fractions and normalized mass fractions of the RACM species.

| RACM species | RACM species molar weight (g $mol^{-1}$) | Corresponding species in the McDonald et al. inventory | Estimated indoor emission fraction | Normalized mass fraction |
|---|---|---|---|---|
| HC3 | 44 | Straight-chain and branched alkanes up to C5 | 17% | 4.2% |
| HC8 | 114 | C6 and higher straight-chain and branched alkanes and all cycloalkanes | 58% | 22.7% |
| TOL | 92 | toluene | 37% | 1.5% |
| XYL | 106 | All higher aromatics | 14% | 1.3% |
| LIM | 136 | All alkenes/cycloalkenes | 93% | 9.5% |
| $CH_3CH_2OH$ | 46 | ethanol | 87% | 27.3% |
| iPrOH | 60 | i-propyl alcohol | 91% | 16.8% |
| KET | 72 | All other oxygenated VOCs | 22% | 16.8% |

**Table S3.** Concentrations of the major (types of) species in the irradiated and unirradiated zones and ratios between them in the GUV254 cases at different ventilation rates.

| GUV254 cases | Species (concentration unit) | Concentration in the irradiated zone | Concentration in the unirradiated zone | Ratio between the concentrations in the irradiated and unirradiated zones |
|---|---|---|---|---|
| Low ventilation | SARS-CoV-2 (quanta) | 1.89E-01 | 6.14E-01 | 0.31 |
| | $O_3$ (molecules $cm^{-3}$) | 9.37E+10 | 9.29E+10 | 1.01 |
| | OH (molecules $cm^{-3}$) | 4.87E+04 | 2.33E+04 | 2.09 |
| | $HO_2$ (molecules $cm^{-3}$) | 8.00E+08 | 7.52E+08 | 1.06 |
| | NO (molecules $cm^{-3}$) | 6.90E+08 | 6.93E+08 | 1.00 |
| | Aldehydes except formaldehyde (molecules $cm^{-3}$) | 4.11E+10 | 4.11E+10 | 1.00 |
| | Ketones except acetone (molecules $cm^{-3}$) | 2.26E+12 | 2.26E+12 | 1.00 |
| | Peroxides (molecules $cm^{-3}$) | 1.21E+11 | 1.21E+11 | 1.00 |
| | Organic nitrates (molecules $cm^{-3}$) | 4.70E+10 | 4.67E+10 | 1.01 |
| | Alkylperoxys (molecules $cm^{-3}$) | 1.63E+09 | 1.18E+09 | 1.39 |
| | Acylperoxys (molecules $cm^{-3}$) | 4.78E+06 | 8.48E+04 | 56.36 |
| | SOA (μg $m^{-3}$) | 9.59E+00 | 9.59E+00 | 1.00 |
| Medium ventilation | SARS-CoV-2 (quanta) | 1.73E-01 | 5.64E-01 | 0.31 |
| | $O_3$ (molecules $cm^{-3}$) | 5.02E+11 | 4.98E+11 | 1.01 |
| | OH (molecules $cm^{-3}$) | 1.56E+06 | 4.75E+05 | 3.27 |
| | $HO_2$ (molecules $cm^{-3}$) | 5.43E+08 | 4.49E+08 | 1.21 |
| | NO (molecules $cm^{-3}$) | 6.68E+09 | 6.76E+09 | 0.99 |
| | Aldehydes except formaldehyde (molecules $cm^{-3}$) | 5.84E+09 | 5.80E+09 | 1.01 |
| | Ketones except acetone | 2.28E+11 | 2.28E+11 | 1.00 |

| | | | | |
|---|---|---|---|---|
| | (molecules $cm^{-3}$) | | | |
| | Peroxides (molecules $cm^{-3}$) | 3.25E+09 | 3.23E+09 | 1.01 |
| | Organic nitrates (molecules $cm^{-3}$) | 3.36E+09 | 3.32E+09 | 1.01 |
| | Alkylperoxys (molecules $cm^{-3}$) | 4.42E+08 | 3.00E+08 | 1.48 |
| | Acylperoxys (molecules $cm^{-3}$) | 5.88E+05 | 1.10E+05 | 5.37 |
| | SOA (μg $m^{-3}$) | 8.05E-01 | 7.98E-01 | 1.01 |
| High ventilation | SARS-CoV-2 (quanta) | 1.45E-01 | 4.77E-01 | 0.30 |
| | $O_3$ (molecules $cm^{-3}$) | 7.08E+11 | 7.02E+11 | 1.01 |
| | OH (molecules $cm^{-3}$) | 4.74E+06 | 7.97E+05 | 5.95 |
| | $HO_2$ (molecules $cm^{-3}$) | 2.27E+08 | 1.26E+08 | 1.80 |
| | NO (molecules $cm^{-3}$) | 1.78E+10 | 1.80E+10 | 0.99 |
| | Aldehydes except formaldehyde (molecules $cm^{-3}$) | 1.27E+09 | 1.23E+09 | 1.03 |
| | Ketones except acetone (molecules $cm^{-3}$) | 7.61E+10 | 7.61E+10 | 1.00 |
| | Peroxides (molecules $cm^{-3}$) | 1.19E+08 | 1.12E+08 | 1.07 |
| | Organic nitrates (molecules $cm^{-3}$) | 8.99E+08 | 8.59E+08 | 1.05 |
| | Alkylperoxys (molecules $cm^{-3}$) | 1.96E+08 | 8.08E+07 | 2.42 |
| | Acylperoxys (molecules $cm^{-3}$) | 2.31E+05 | 5.61E+04 | 4.11 |
| | SOA (μg $m^{-3}$) | 1.75E-01 | 1.67E-01 | 1.05 |

**Table S4.** Mass yields of secondary organic aerosol produced through several volatile organic compound oxidation pathways.

| VOC + oxidant | Mass yield | Source |
|---|---|---|
| LIM + $O_3$ | 20% | Estimated based on ref 23 |
| LIM + OH | 13% | Ref 6 |
| HC8 + OH | 10% | Rough average of yields for corresponding species in the inventory of ref 6 |
| KET + OH | 5% | Rough average of yields for corresponding species in the inventory of ref 6 |
| TOL + OH | 9% | Ref 6 |
| XYL + OH | 4.9% | Ref 6 |

(a)

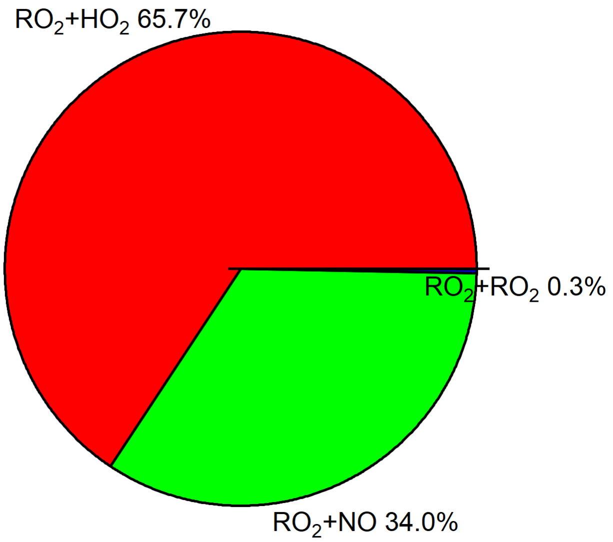
RO2+HO2 65.7%
RO2+RO2 0.3%
RO2+NO 34.0%

(b)

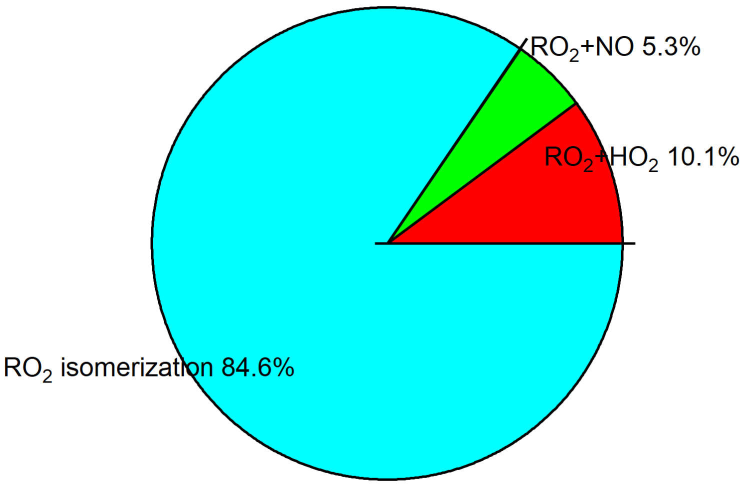
RO2+NO 5.3%
RO2+HO2 10.1%
RO2 isomerization 84.6%

(c)

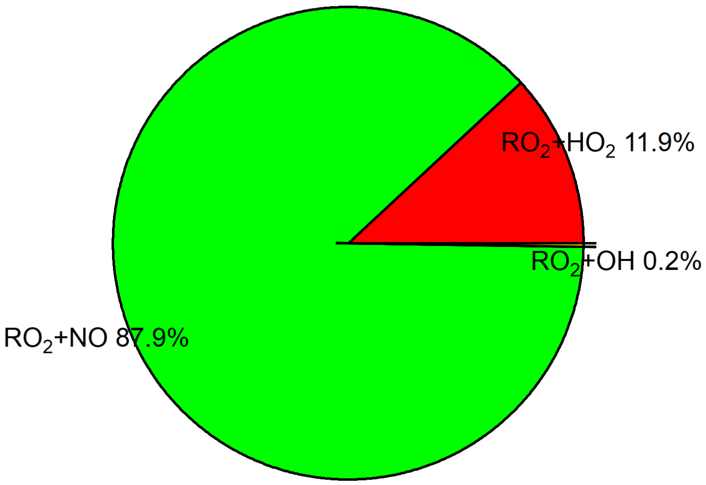

(d)

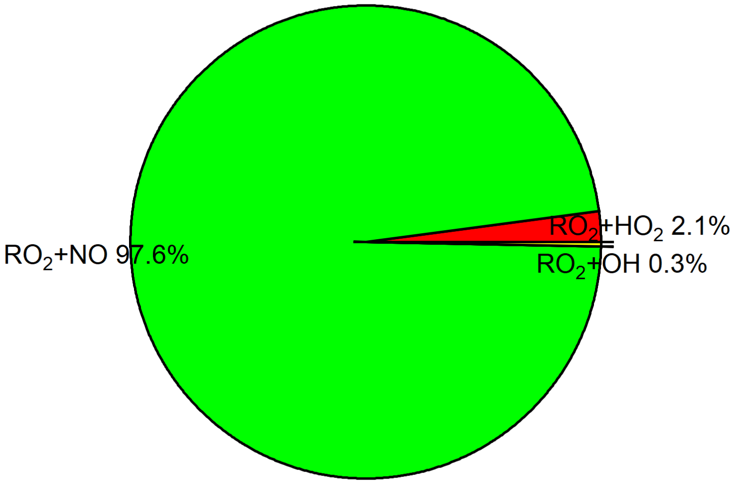

**Figure S1.** Alkyl $RO_2$ fate estimated per Peng et al.:[24] (a) bimolecular fate and (b) fate including unimolecular isomerization in the low-ventilation GUV254 case, and bimolecular fate in the (c) medium- and (d) high-ventilation GUV254 cases. $RO_2$ unimolecular isomerization rate coefficient is assumed to be 0.1 $s^{-1}$, typical for oxygenated VOCs.[25] "$RO_2$+$RO_2$" denotes the reaction between an alkyl $RO_2$ and an acyl $RO_2$, with a rate coefficient assumed to be $1 \times 10^{-11}$ $cm^3$ $molecule^{-1}$ $s^{-1}$.[26]
